# Efficacy Estimates for Various COVID-19 Vaccines: What we Know from the Literature and Reports

**DOI:** 10.1101/2021.05.20.21257461

**Authors:** Julia Shapiro, Natalie E. Dean, Zachary J. Madewell, Yang Yang, M.Elizabeth Halloran, Ira Longini

## Abstract

In this report, we provide summary estimates, from publications and reports, of vaccine efficacy (VE) for the COVID-19 vaccines that are being rolled out on a global scale. We find that, on average, the efficacy against any disease with infection is 85% (95% CI: 71 - 93%) after a fully course of vaccination. The VE against severe disease, hospitalization or death averages close to 100%. The average VE against infection, regardless of symptoms, is 84% (95% CI: 70 - 91%). We also find that the average VE against transmission to others for infected vaccinated people is 48% (95% CI: 45 - 52%). Finally, we prove summary estimates of the VE against any disease with infection for some of the variants of concern (VOC). The average VE for the VOC *γ* (P1) is 61% (95% CI: 43 - 73%). The average VE for the VOC *α* (B.1.1.7), *β* (B.1.351), and *δ* (B.1.617.2) after dose 1 are 48% (95% CI: 44 - 51%), 35% (95% CI: -11 - 62%), and 33% (95% CI: 21 - 43%), respectively. The average VE for the VOC *α* (B.1.1.7), *β* (B.1.351), and *δ* (B.1.617.2) after dose 2 are 85% (95% CI: 25 - 97%), 57% (95% CI: 14 - 78%), and 78% (95% CI: 28 - 93%), respectively.

## Introduction

In this report, we summarize estimates of vaccine efficacy (VE) for the COVID-19 vaccines that are being rolled out on local and global scales. This includes the Pfizer, Moderna, Johnson & Johnson, AstraZeneca, Sputnik, Novavax, Sinovac, and Sinopharm vaccines. VE estimates are taken from journal articles and media reports for the vaccines that have gone through double-blinded, placebo-controlled, phase III vaccine trials, as well as observational studies. Some of the estimates are based on rigorous, preplanned statistical analyses from double-blinded, placebo-controlled trials, while others are extracted from observational studies with different levels of control. These studies are reported from a variety of sources including publications, reports, and sometimes press releases. Because of this, we do not carry out a formal meta analysis. In all cases, we try to extract estimates for one or more of the triplet of vaccine efficacy parameters (*V E*_*S*_, *V E*_*P*_, *V E*_*I*_) [1], where *V E*_*S*_ is VE against infection; *V E*_*P*_ is VE against disease, given infection; and *V E*_*I*_ is VE against transmission to others, given infection. A fourth parameter, *V E*_*SP*_, which is VE against disease and infection, tends to be available from vaccine trials, and it is the usual primary outcome for those trials (i.e., cases of disease that are confirmed infections). The *V E*_*SP*_ is a function of both the *V E*_*S*_ and *V E*_*P*_. If we believe in a multiplicative and independent relationship, then *V E*_*SP*_ = 1–(1—*V E*_*S*_)(1 −*V E*_*P*_). Thus, if we have two of these VE’s, we can always calculate the third.

In the material that follows, we give estimates of these VE’s as a function of time when protection is believed to begin to occur after the first and second dose for two-dose vaccines, and after the first dose for one-dose vaccines. We also provide *V E*_*SP*_ estimates for protection against the variants of concern (VOC) *γ* (P1), *α* (B.1.1.7), *β* (B.1.351), and *δ* (B.1.617.2). The methods for creating the forest plots are given in the Appendix. The supporting tables for the analysis are also given in the Appendix. Not all estimates described in the tables are given in the figures, as we have tried to extract the essential information without getting lost in too much detail. However, virtually all the complete information is given in the Appendix tables.

## Results

We first consider VE for the original wild type viruses. Figure 1 (Table A1) give the estimates of the *V E*_*SP*_ after the second dose for two-dose vaccines. All the estimates are from double-blinded, placebo-controlled vaccine trials. With the exception of the Sinovac vaccine, they are all over 80%, with a summary estimate of 85% (95% CI: 71 - 93%). The Sinovac *V E*_*SP*_ estimate is 51% (95% CI: 36 - 62%).

**Figure 1:**
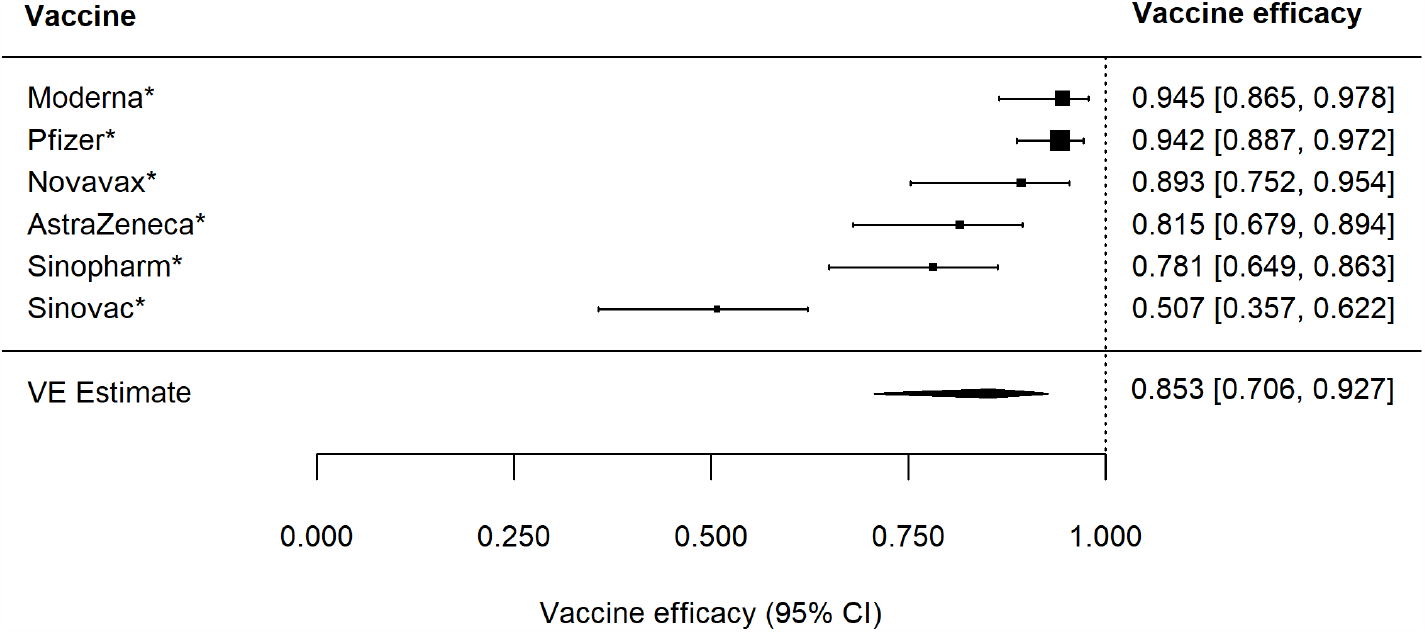
Forest plot of vaccine efficacy to prevent any disease after dose 2, *V E*_***SP***_. * indicates double-blinded, randomized vaccine trial.

The estimated *V E*_*SP*_ after one dose, for both two-dose and one-dose vaccines, is given in Figure 2 (Table A2), where the Johnson & Johnson vaccine is the only one-dose vaccine listed. The estimates are generally almost as high as protection after one dose, with summary estimated of 82% (95% CI: 72 - 88%).

**Figure 2:**
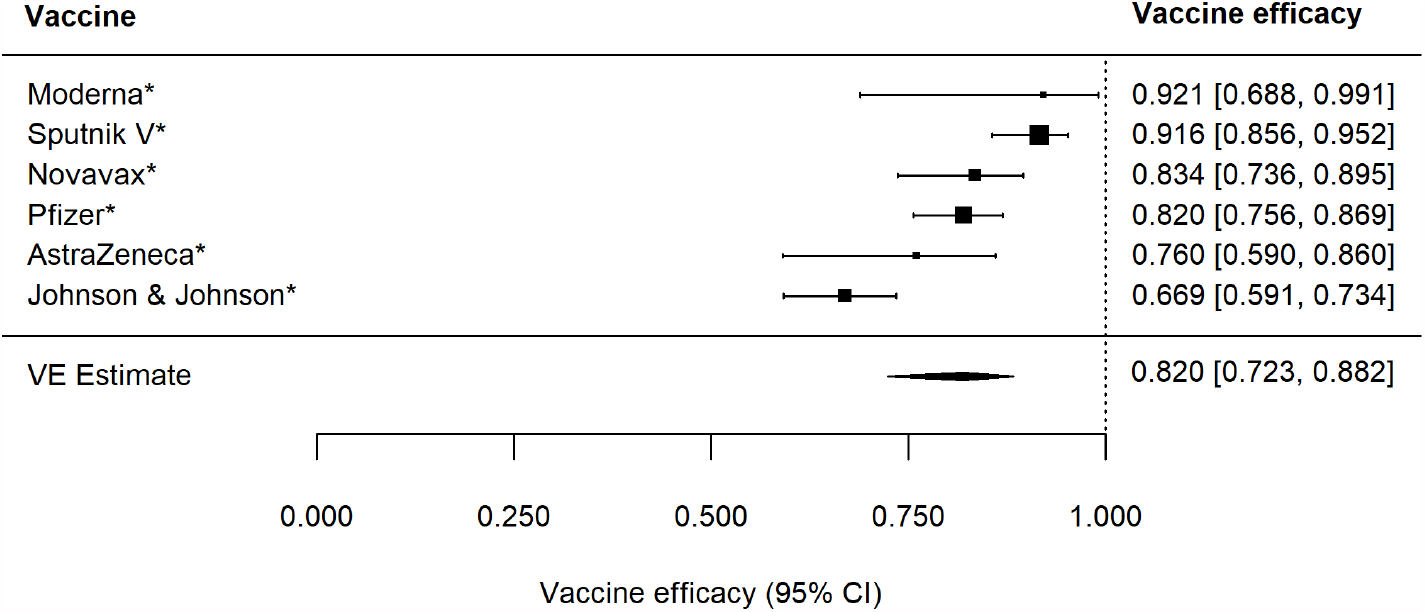
Forest plot of vaccine efficacy to prevent any disease after dose 1, *V E*_***SP***_. * indicates double-blinded, randomized vaccine trial.

Figure 3 (Table A3) give the estimates of the 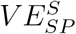 (VE for severe disease with infection) after the second dose for two-dose vaccines. The estimates a very high, and generally close to 100%, with relatively poor precision.

**Figure 3:**
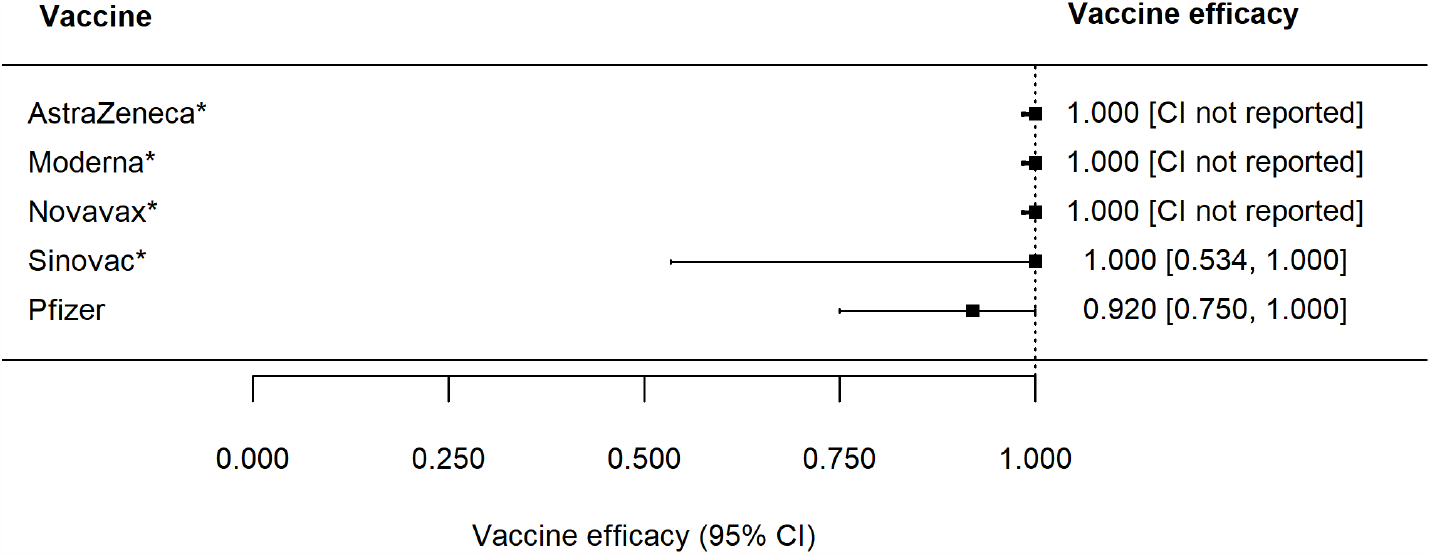
Forest plot of vaccine efficacy to prevent severe disease after dose 2, 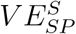. * indicates double-blinded, randomized vaccine trial.

Figure 4 (Table A4) give the estimates of the 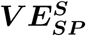 (VE for severe disease with infection) after the first dose for two-dose vaccines and one dose for the one-dose vaccine. The summary estimated is quite high at 86% (95% CI: 39 - 97%).

**Figure 4:**
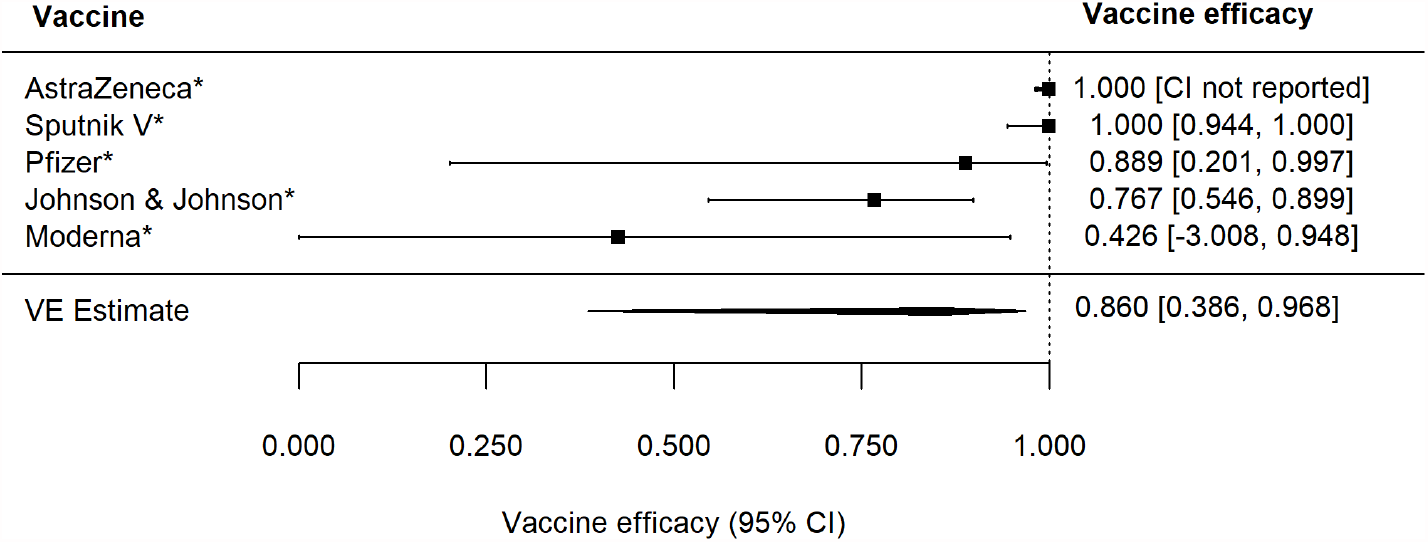
Forest plot of vaccine efficacy to prevent severe disease after dose 1, 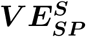. * indicates double-blinded, randomized vaccine trial.

VE against hospitalization and death were quite high, as shown in Figures 5 and 6 (Tables A5 and A6).

**Figure 5:**
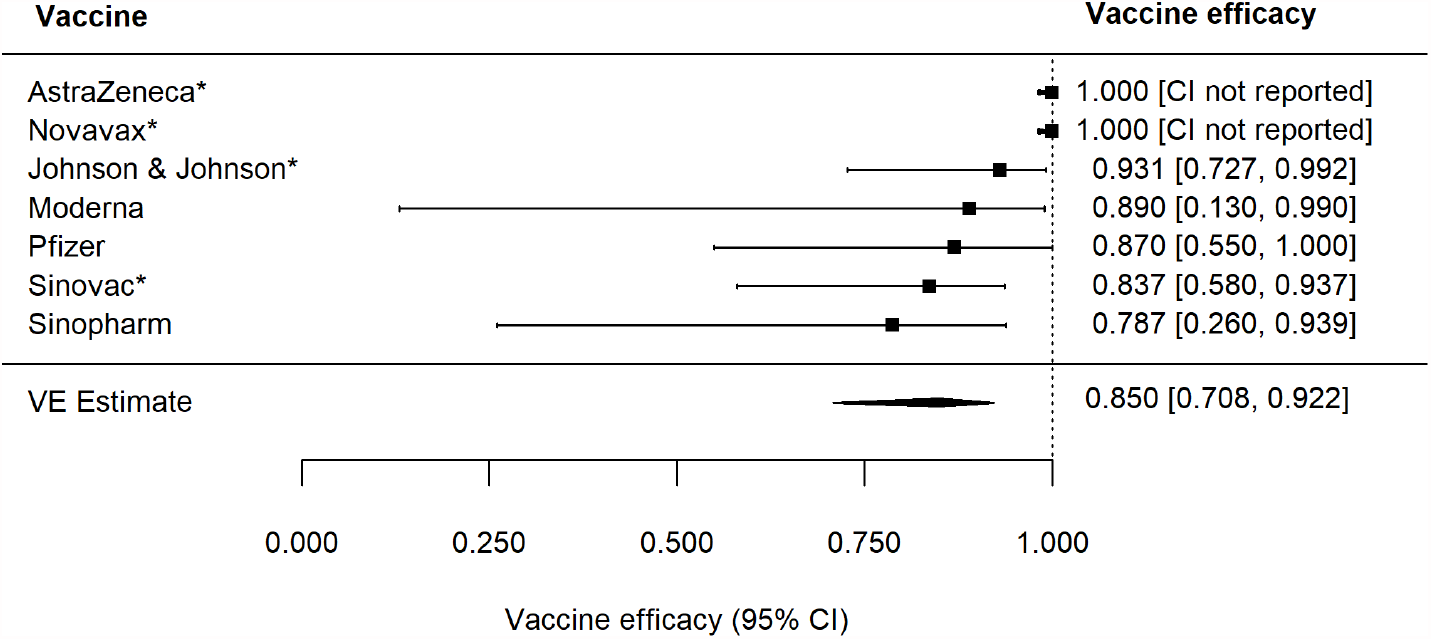
Forest plot of vaccine efficacy to prevent hospitalization, 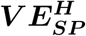. * indicates double-blinded, randomized vaccine trial.

**Figure 6:**
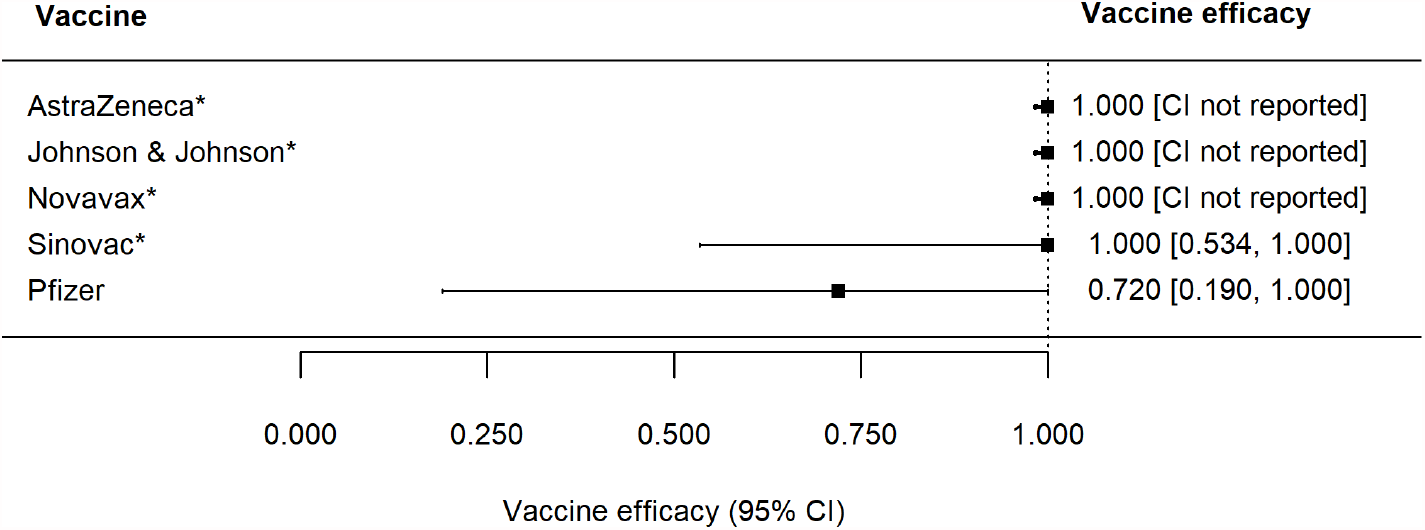
Forest plot of vaccine efficacy to prevent death, 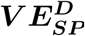. * indicates double-blinded, randomized vaccine trial.

Figure 7 (Table A7) give the estimates of the *V E*_*S*_, i.e., VE against infection. The estimates were quite high, with a summary estimate of 84% (95% CI: 70 - 91%).

**Figure 7:**
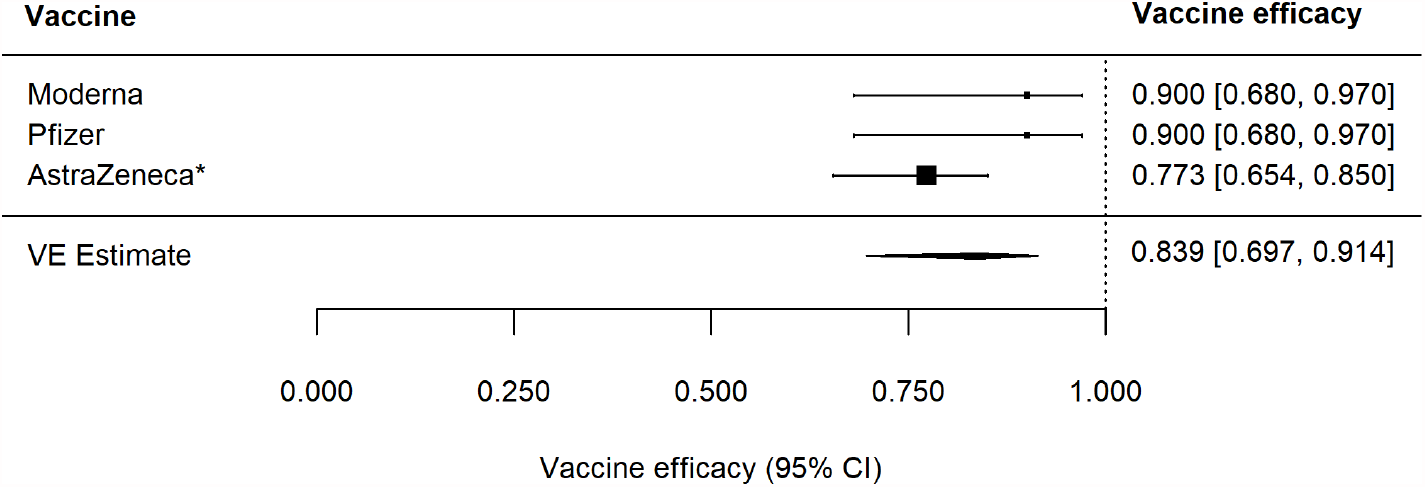
Forest plot of vaccine efficacy to prevent infection, *V E*_***S***_. * indicates double-blinded, randomized vaccine trial.

**Figure 8:**
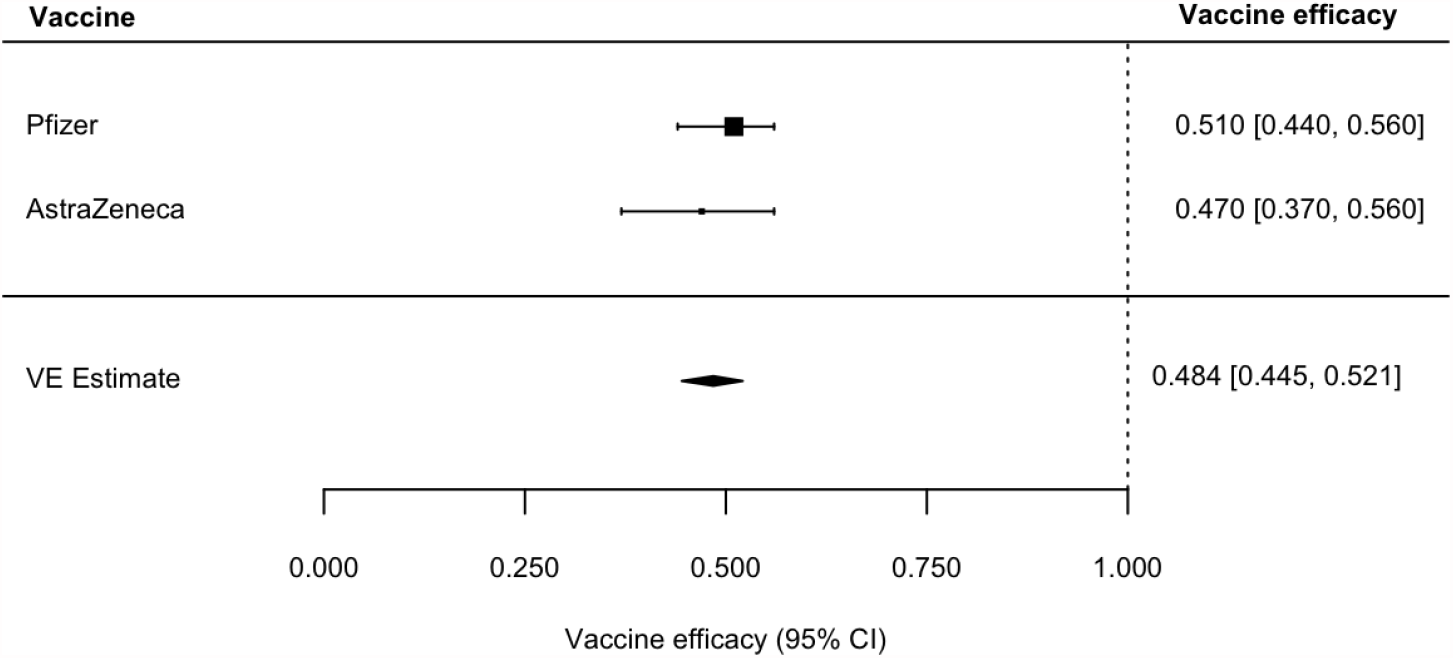
**Forest plot of vaccine efficacy to prevent infectiousness to others, *V E*_*I*_**

(Table A8) give estimates of the *V E*_*I*_, i.e., VE against infectiousness or direct transmission to others. The summary measure is 48% (95% CI: 45 - 52%), indicating the vaccination reduces the transmission to others by 48% when vaccinated people are infected, compared to unvaccinated people who become infected.

Now we consider VE’s for the variants of concern (VOC). Estimates are available for the *V E*_*SP*_, mostly after the first dose for the one dose vaccine and the second dose for the two dose vaccines. These estimates are given in Figures 9-15 (Tables A9-A15).

**Figure 9:**
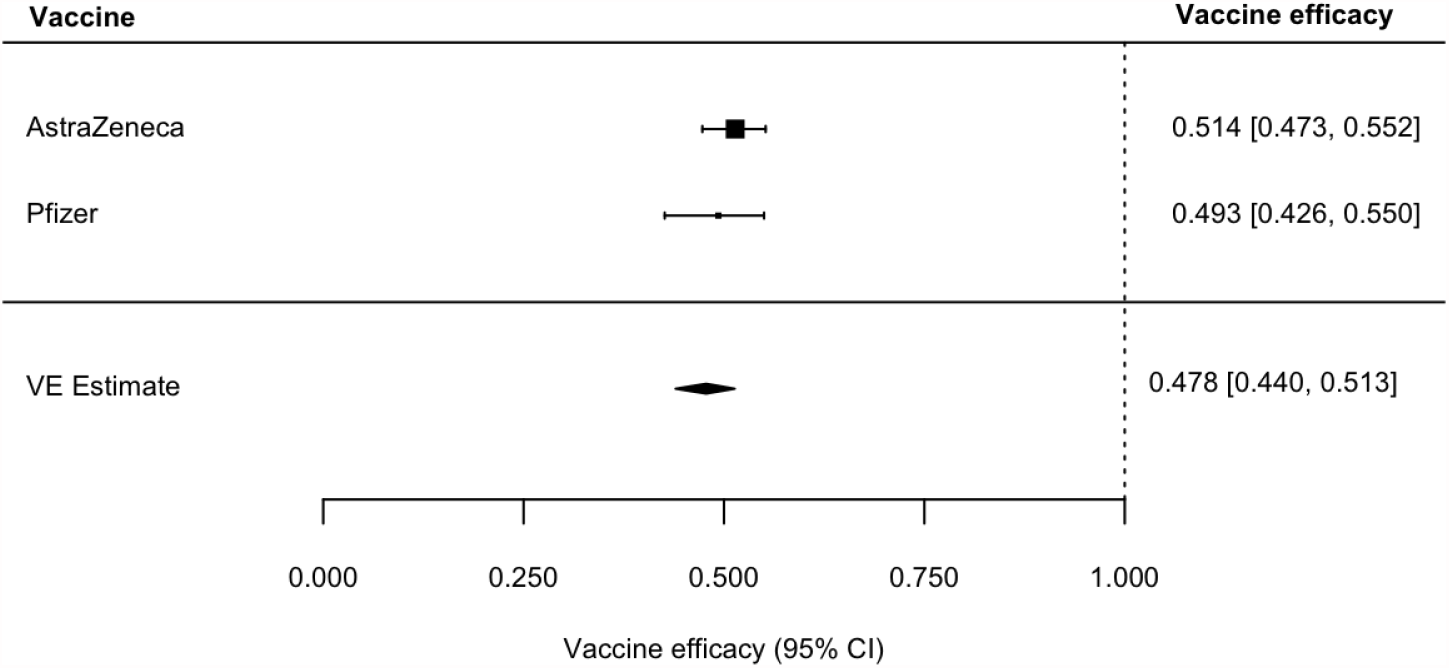
Forest plot of vaccine efficacy against *α* (B.1.1.7) after dose 1.

**Figure 10:**
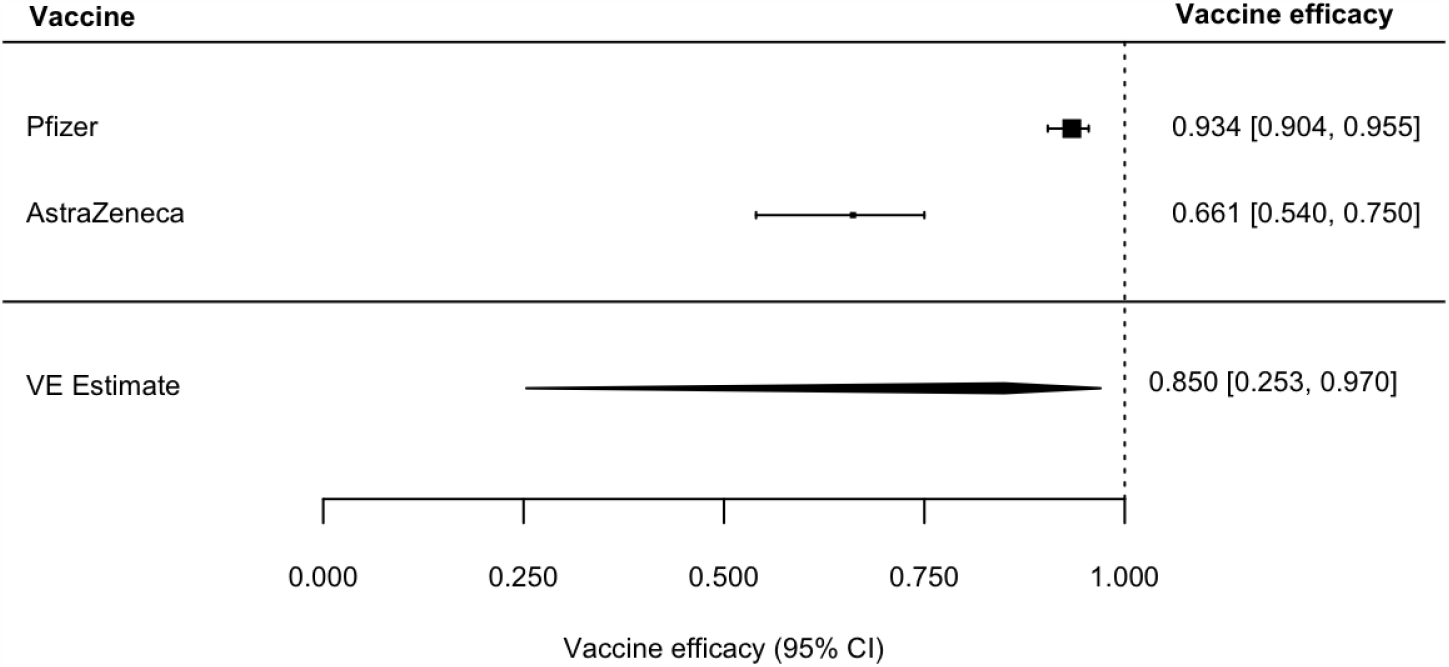
Forest plot of vaccine efficacy against *α* (B.1.1.7) after dose 2.

**Figure 11:**
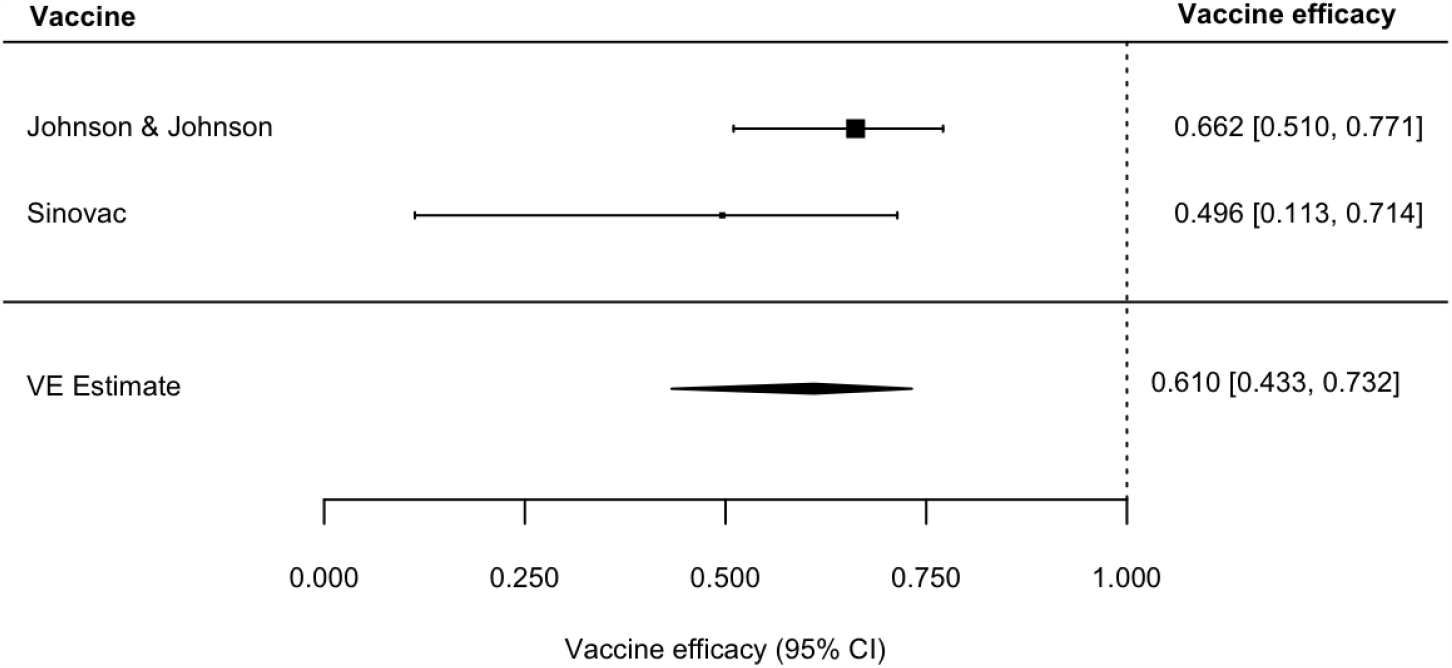
Forest plot of vaccine efficacy against (*γ*) (P.1)

**Figure 12:**
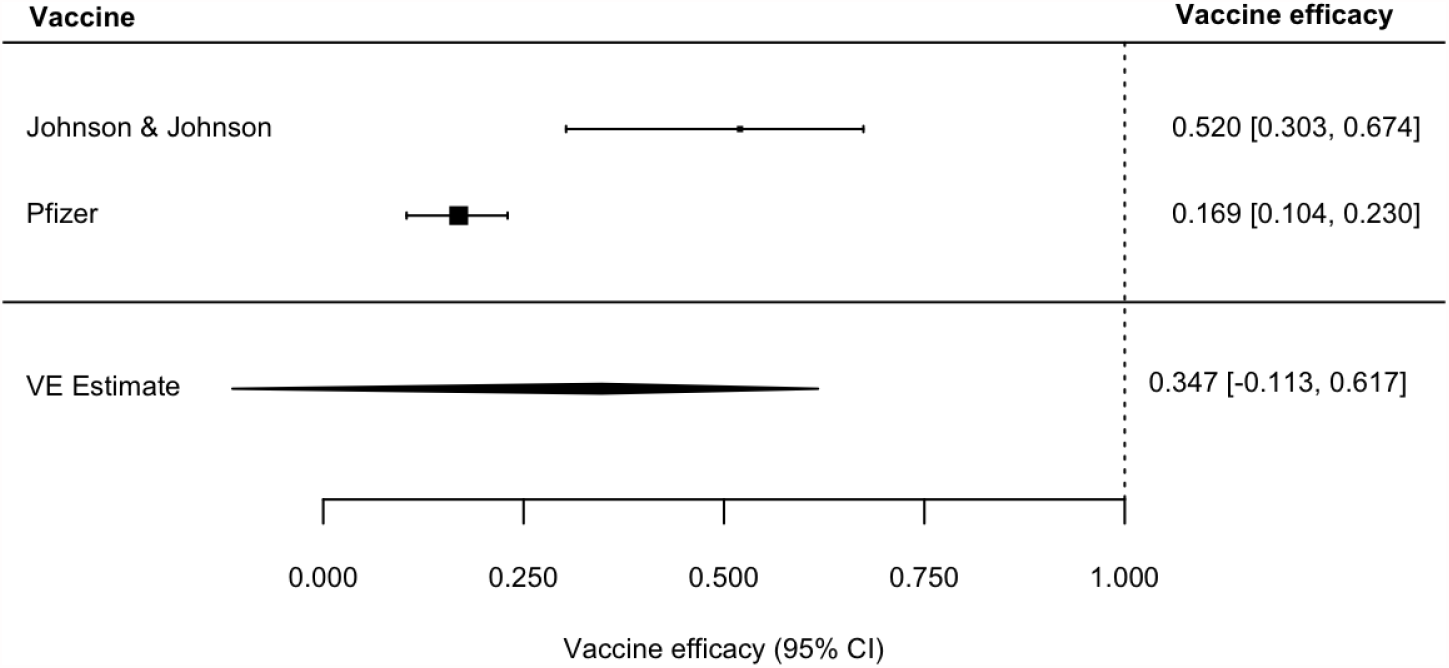
Forest plot of vaccine efficacy against *β* (B.1.351) after dose 1.

**Figure 13:**
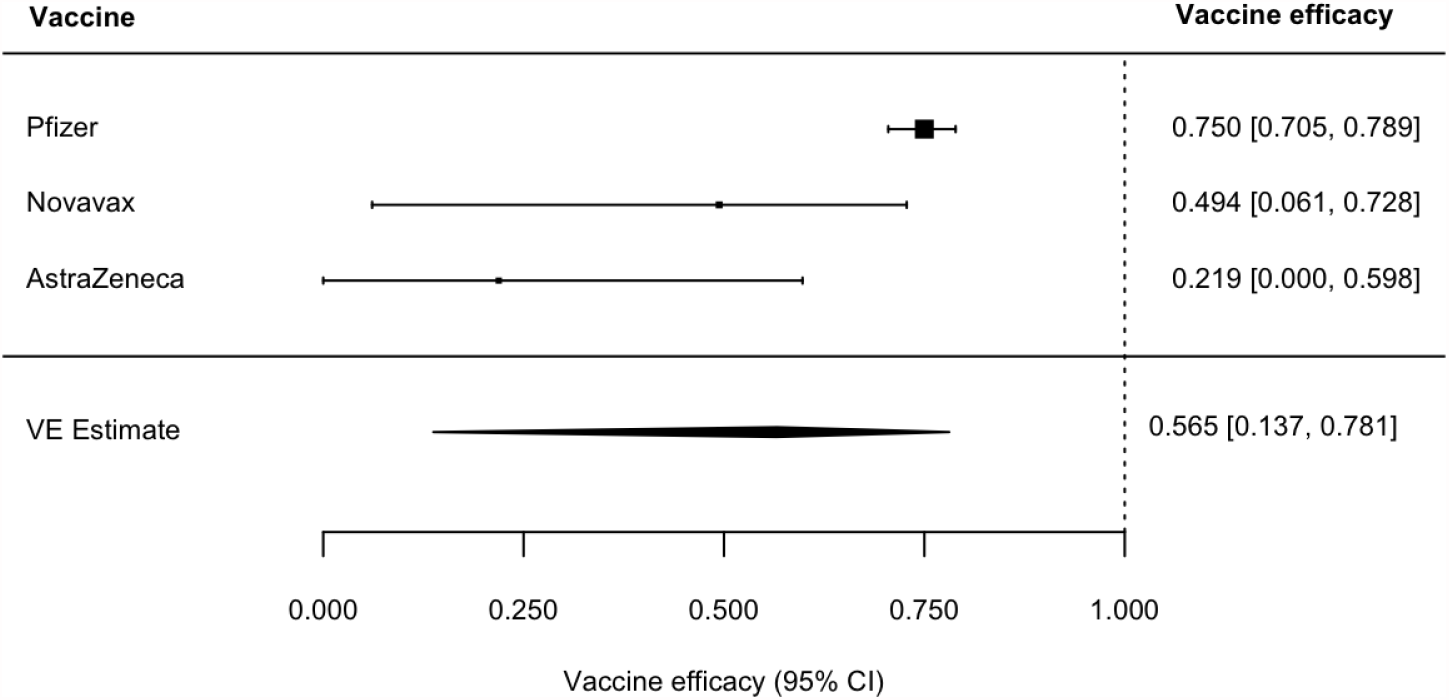
Forest plot of vaccine efficacy against *β* (B.1.351) after dose 2.

**Figure 14:**
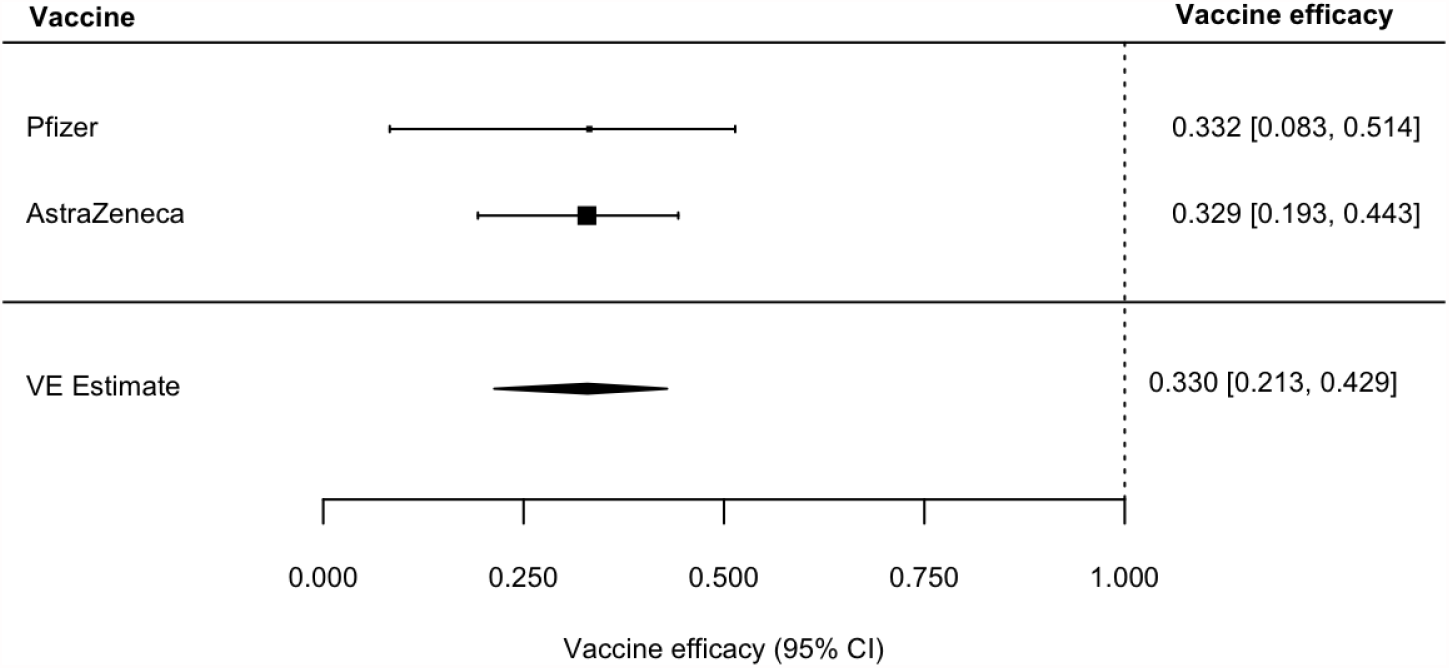
Forest plot of vaccine efficacy against *δ* (B.1.617.2) after dose 1.

**Figure 15:**
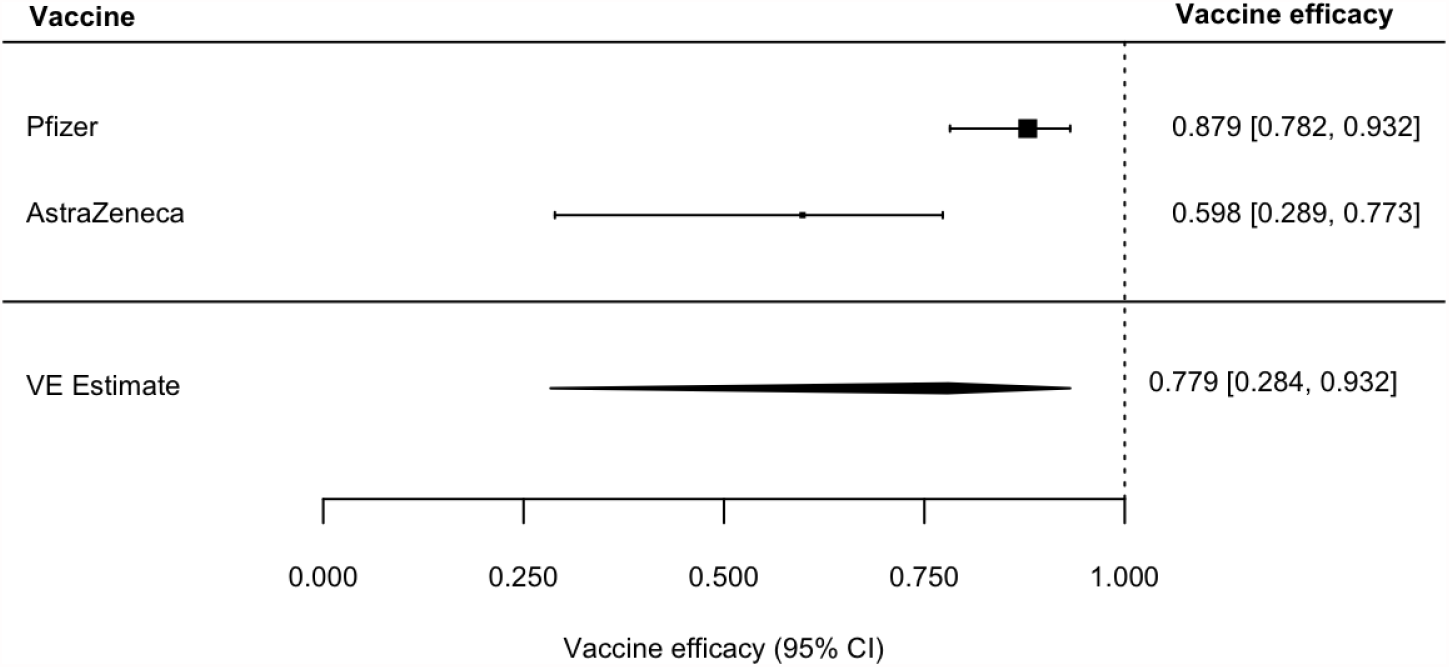
Forest plot of vaccine efficacy against *δ* (B.1.617.2) after dose 2.

For *α* (B.1.1.7), VE is 48% (95% CI: 44 - 51%) after dose 1, and VE is 85% (95% CI: 25 - 97%) after dose 2. The summary estimate for VE after dose 1 is considerably lower than the VE for the wild type virus. In contrast, however, the summary estimate after dose 2 is just somewhat reduced compared to the wild type virus. It is necessary to note that this VOC does not have a mutation that affects immunity, whereas the other VOC’s have mutations that affect immune function.

The average VE for the VOC *γ* (P1) is 61% (95% CI: 43 - 73%). For *β* (B.1.351), VE is 35% (95% CI: -11 - 62%) after dose 1, and VE is 57% (95% CI: 14 - 78%) after dose 2. The summary estimates for VE after dose 1 and dose 2 are considerably lower than the VE’s for the wild type virus.

For *δ* (B.1.617.2), VE is 33% (95% CI: 21 - 43%) after dose 1, and VE is 78% (95% CI: 28 - 93%) after dose 2. The summary estimate for VE after dose 1 is considerably lower than the VE for the wild type virus. In contrast, however, the summary estimate after dose 2 is just somewhat reduced compared to the wild type virus.

## Discussion

We have presented the relevant VE estimates for the COVID-19 vaccines that are being rolled out on a global scale and for which there is sufficient quality data. We provide estimates of VE against disease with confirmed infection, infection, and transmission to others. The VE estimates against disease are stratified by disease severity, hospitalization and death. We have also provided VE estimates for three of the VOC.

These estimates should be useful for constructing mathematical models for vaccination impact and for making policy decisions involving vaccination. We plan to keep updating this report as more information becomes available.

## Methods

For each vaccine efficacy measure (e.g., severe disease, infection), we first obtained log odds ratios and corresponding sampling variances from each vaccine efficacy estimate and 95% confidence interval (CI). We then fit random-effects models to these data to estimate average log odds ratios, which we back-transformed to obtain VE summary estimates and 95% CIs. All analyses were done in R version 4.0.2 using the package metafor (R Project for Statistical Computing) [2, 3].

## Data Availability

We agree to make data used to compile this paper available

## Appendix

Here we give the details about the studies and data that are summarized in the figures. **Note:** ^*\**^ indicates that the VE estimate is based on double-blinded, randomized trials.

**Table 1:** Vaccine efficacy to prevent any disease after dose 2, *V E*_***SP***_.

| Company | Efficacy%, (95% CI),<br>time frame of estimate | References |
| --- | --- | --- |
| Moderna | 94.5*, (86.5, 97.8), 14 or<br>more days after dose 2 | [4] |
| Pfizer | 94.2*, (88.7, 97.2), 14 or<br>more days after dose 2 | [5] |
| Johnson & Johnson | One dose vaccine | n/a |
| AstraZeneca | 81.5*, (67.9, 89.4), 14 or<br>more days after dose 2 | [6] |
| Novavax | 89.3*, (75.2, 95.4), 7 or<br>more days after dose 2 | [7] |
| Sputnik V | Not reported | n/a |
| Sinovac | 50.7*, (35.7, 62.2), time<br>frame not reported | [8] |
| Sinopharm | 78.1*, (64.9, 86.3), median<br>follow-up time 112 days | [9] |

**Table 2:** Vaccine efficacy to prevent any disease after dose 1, *V E*_***SP***_.

| Company | Efficacy%, (95% CI),<br>time frame of estimate | References |
| --- | --- | --- |
| Moderna | 92.1*, (68.8, 99.1), more<br>than 14 days after dose 1 | [4] |
| Pfizer | 82.0*, (75.6, 86.9), after<br>dose 1 | [5] |
|  | 57.0, (50.0, 60.0), 14-20<br>day period after dose 1 | [10] |
| Johnson & Johnson | 66.9*, (59.1, 73.4), 14 or<br>more days after vaccination | [11,12] |
|  | 66.5*, (55.5, 75.1), 28 or<br>more days after vaccination | [11,12] |
| AstraZeneca | 76.0*, (59.0, 86.0), 22-90<br>day period after dose 1 | [7] |
| Novavax | 83.4*, (73.6, 89.5), 14 or<br>more days after dose 1 | [13] |
| Sputnik V | 91.6*, (85.6, 95.2), 21 days<br>after dose 1 | [14] |
| Sinovac | Not reported | n/a |
| Sinopharm | Not reported | n/a |

**Table 3:** Vaccine efficacy to prevent severe disease after dose 2, 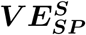.

| Company | Efficacy%, (95% CI),<br>time frame of estimate | References |
| --- | --- | --- |
| Moderna | 100.0*, (CI not reported),<br>14 or more days after dose<br>2 | [4] |
| Pfizer | 92.0, (75.0, 100.0), 7 or<br>more days after dose 2 | [10] |
| Johnson & Johnson | One dose vaccine | n/a |
| AstraZeneca | 100.0*, (CI not reported),<br>time frame not reported | [7] |
| Novavax | 100.0*, (CI not reported),<br>time frame not reported | [7] |
| Sputnik V | Not reported | n/a |
| Sinovac | 100.0*, (53.4, 100.0)<sup>a</sup>, time<br>frame not reported | [8] |
| Sinopharm | Not reported | n/a |

**Table 4:** Vaccine efficacy to prevent severe disease after dose 1, 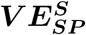.

| Company | Efficacy%, (95% CI),<br>time frame of estimate | References |
| --- | --- | --- |
| Moderna | 42.6*, (-300.8, 94.8), 14 or<br>more days after dose 1 | [4] |
| Pfizer | 88.9*, (20.1, 99.7), after<br>dose 1 | [5] |
|  | 62.0, (39.0, 80.0), 14-20<br>day period after dose 1 | [10] |
| Johnson & Johnson | 76.7*, (54.6, 89.9), 14 or<br>more days after vaccination | [11, 12] |
|  | 85.4*, (54.2, 96.9), 28 or<br>more days after vaccination | [11, 12] |
| AstraZeneca | 100.0*, (CI not reported),<br>more than 22 days after<br>dose 1 | [15] |
| Novavax | Not reported | n/a |
| Sputnik V | 100.0*, (94.4, 100.0), 21 or<br>more days after dose 1 | [16] |
| Sinovac | Not reported | n/a |
| Sinopharm | Not reported | n/a |

**Table 5:** Vaccine efficacy to prevent hospitalization, 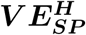.

| Company | Efficacy%, (95% CI),<br>time frame of estimate | References |
| --- | --- | --- |
| Moderna | 89.0, (13.0, 99.0), time<br>frame not reported | [17] |
| Pfizer | 87.0, (55.0, 100.0), 7 or<br>more days after dose 2 | [10] |
|  | 74.0, (56.0, 86.0), 14-20<br>days after dose 1 | [10] |
|  | 91.0, (85.0, 94.0), 28-34<br>days after a single dose | [18] |
| Johnson & Johnson | 93.1*, (72.7, 99.2), 14 or<br>more days after vaccination | [11, 12] |
|  | 100.0*, (74.3, 100.0), 28 or<br>more days after vaccination | [11, 12] |
| AstraZeneca | 100.0*, (CI not reported),<br>more than 22 days after<br>dose 1 | [15] |
|  | 88.0, (75.0, 94.0), 28-34<br>days after a single dose | [18] |
| Novavax | 100.0*, (CI not reported),<br>time frame not reported | [13] |
| Sputnik V | Not reported | n/a |
| Sinovac | 83.7*, (58.0, 93.7), time<br>frame not reported | [8] |
| Sinopharm | 78.7*, (26.0, 93.9), median<br>follow-up time 112 days | [9] |

**Table 6:**
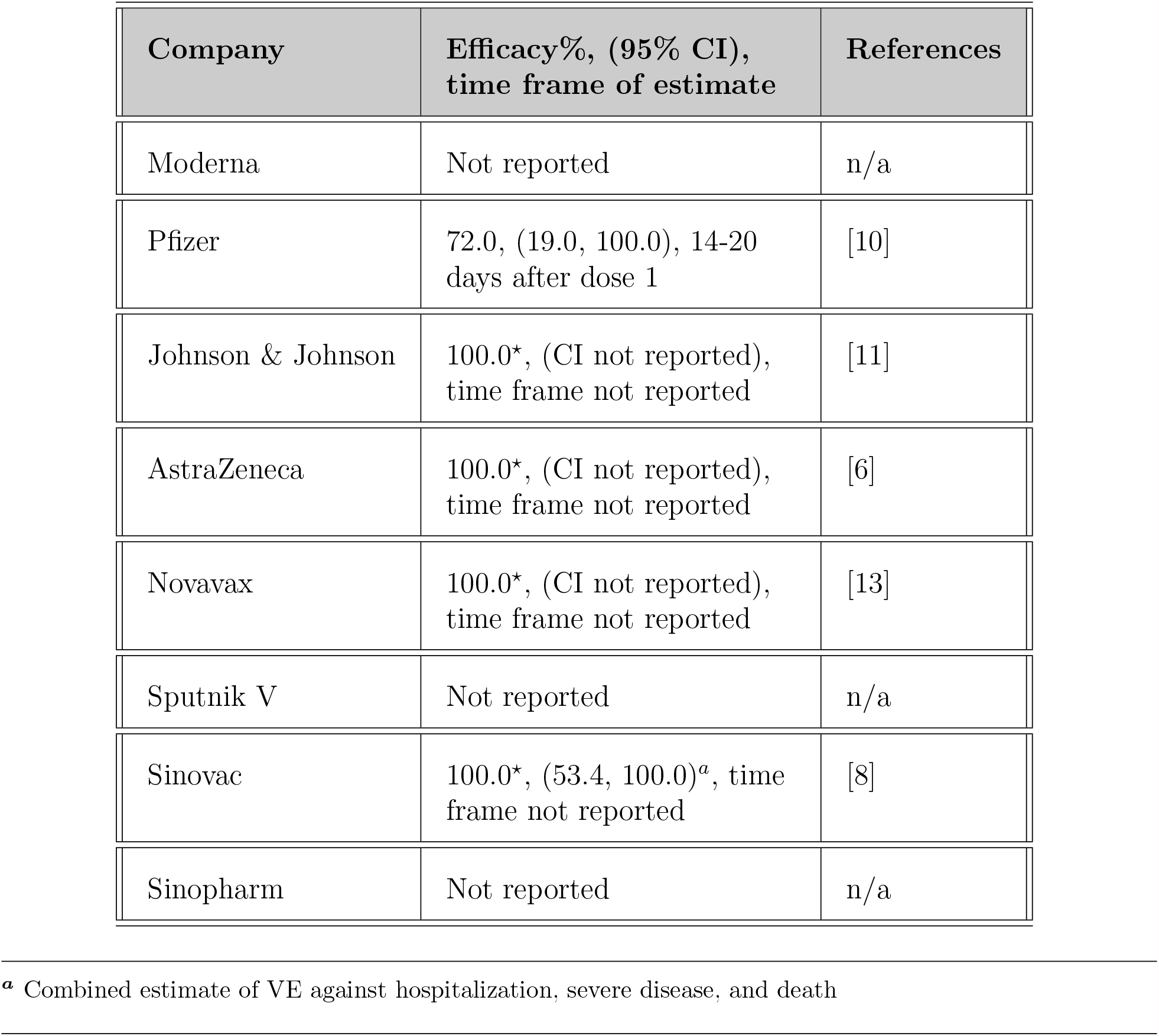
Vaccine efficacy to prevent death, 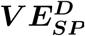.

| Company | Efficacy%, (95% CI),<br>time frame of estimate | References |
| --- | --- | --- |
| Moderna | Not reported | n/a |
| Pfizer | 72.0, (19.0, 100.0), 14-20<br>days after dose 1 | [10] |
| Johnson & Johnson | 100.0*, (CI not reported),<br>time frame not reported | [11] |
| AstraZeneca | 100.0*, (CI not reported),<br>time frame not reported | [6] |
| Novavax | 100.0*, (CI not reported),<br>time frame not reported | [13] |
| Sputnik V | Not reported | n/a |
| Sinovac | 100.0*, (53.4, 100.0)<sup>a</sup>, time<br>frame not reported | [8] |
| Sinopharm | Not reported | n/a |

**Table 7:**
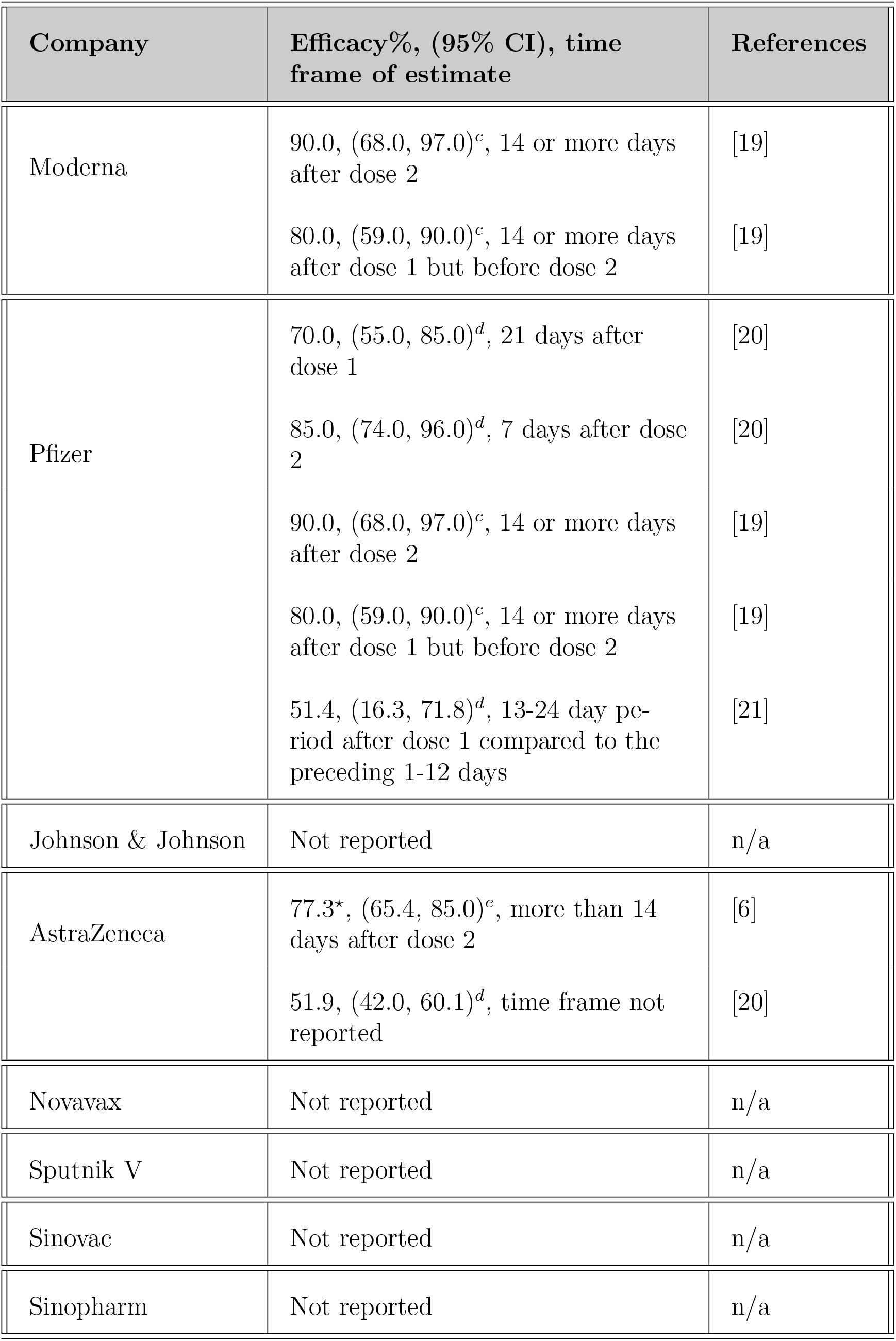
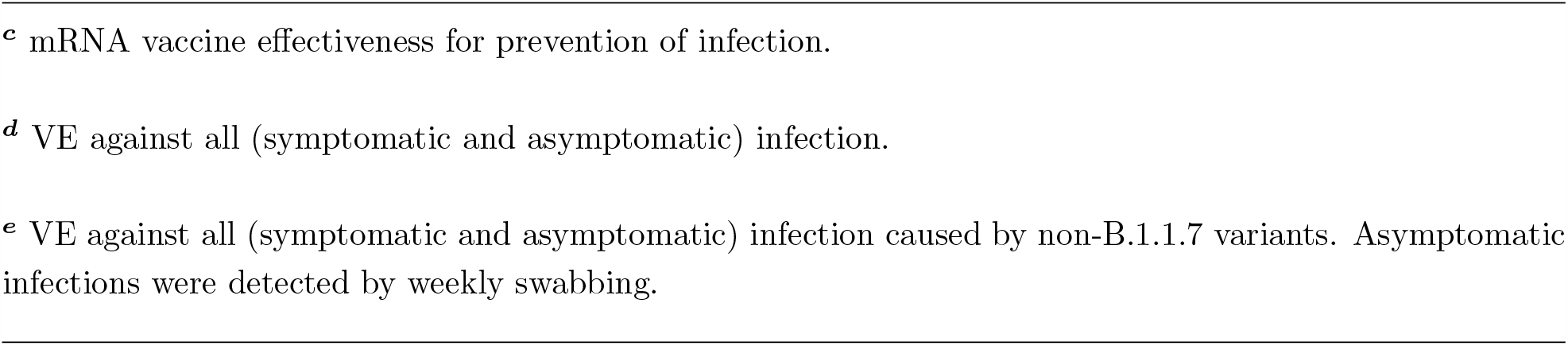
Vaccine efficacy to prevent infection, *V E*_***S***_.

| Company | Efficacy%, (95% CI), time frame of estimate | References |
| --- | --- | --- |
| Moderna | 90.0, (68.0, 97.0)<sup>c</sup>, 14 or more days after dose 2 | [19] |
|  | 80.0, (59.0, 90.0)<sup>c</sup>, 14 or more days after dose 1 but before dose 2 | [19] |
| Pfizer | 70.0, (55.0, 85.0)<sup>d</sup>, 21 days after dose 1 | [20] |
|  | 85.0, (74.0, 96.0)<sup>d</sup>, 7 days after dose 2 | [20] |
|  | 90.0, (68.0, 97.0)<sup>c</sup>, 14 or more days after dose 2 | [19] |
|  | 80.0, (59.0, 90.0)<sup>c</sup>, 14 or more days after dose 1 but before dose 2 | [19] |
|  | 51.4, (16.3, 71.8)<sup>d</sup>, 13-24 day period after dose 1 compared to the preceding 1-12 days | [21] |
| Johnson & Johnson | Not reported | n/a |
| AstraZeneca | 77.3*, (65.4, 85.0)<sup>e</sup>, more than 14 days after dose 2 | [6] |
|  | 51.9, (42.0, 60.1)<sup>d</sup>, time frame not reported | [20] |
| Novavax | Not reported | n/a |
| Sputnik V | Not reported | n/a |
| Sinovac | Not reported | n/a |
| Sinopharm | Not reported | n/a |

**Table 8:**
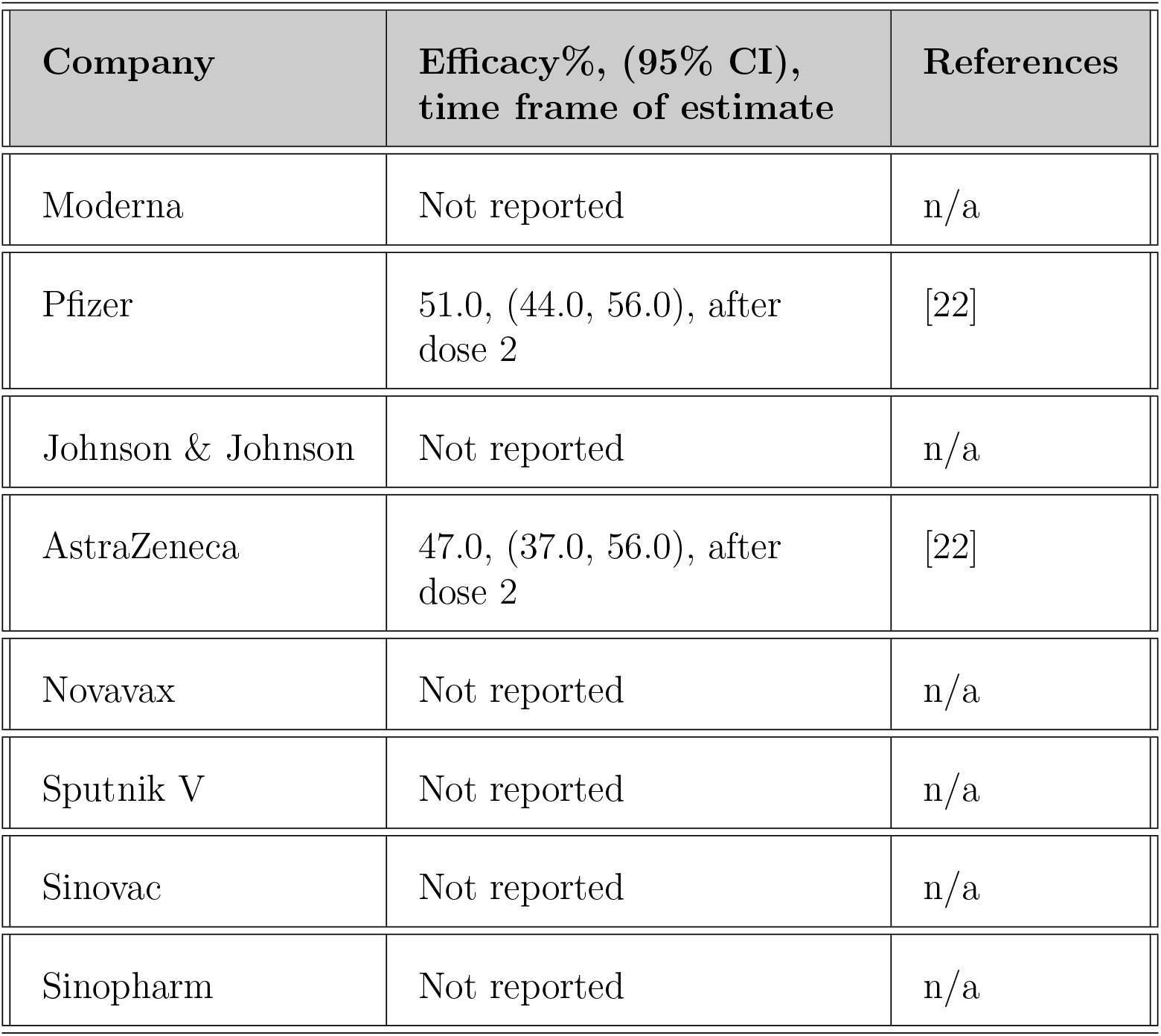
Vaccine efficacy to prevent infectiousness to others, *V E*_***I***_.

**Table 9:**
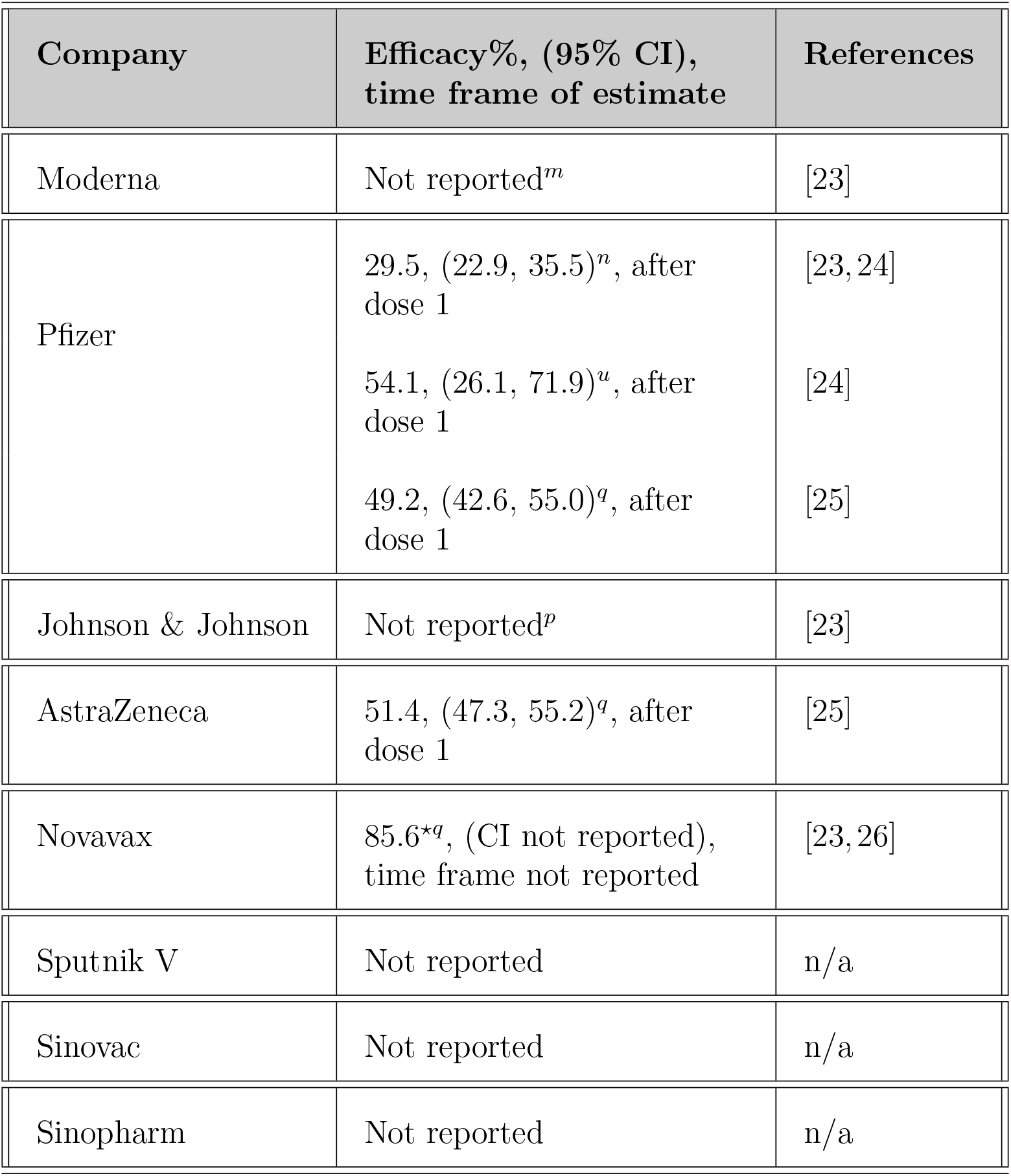
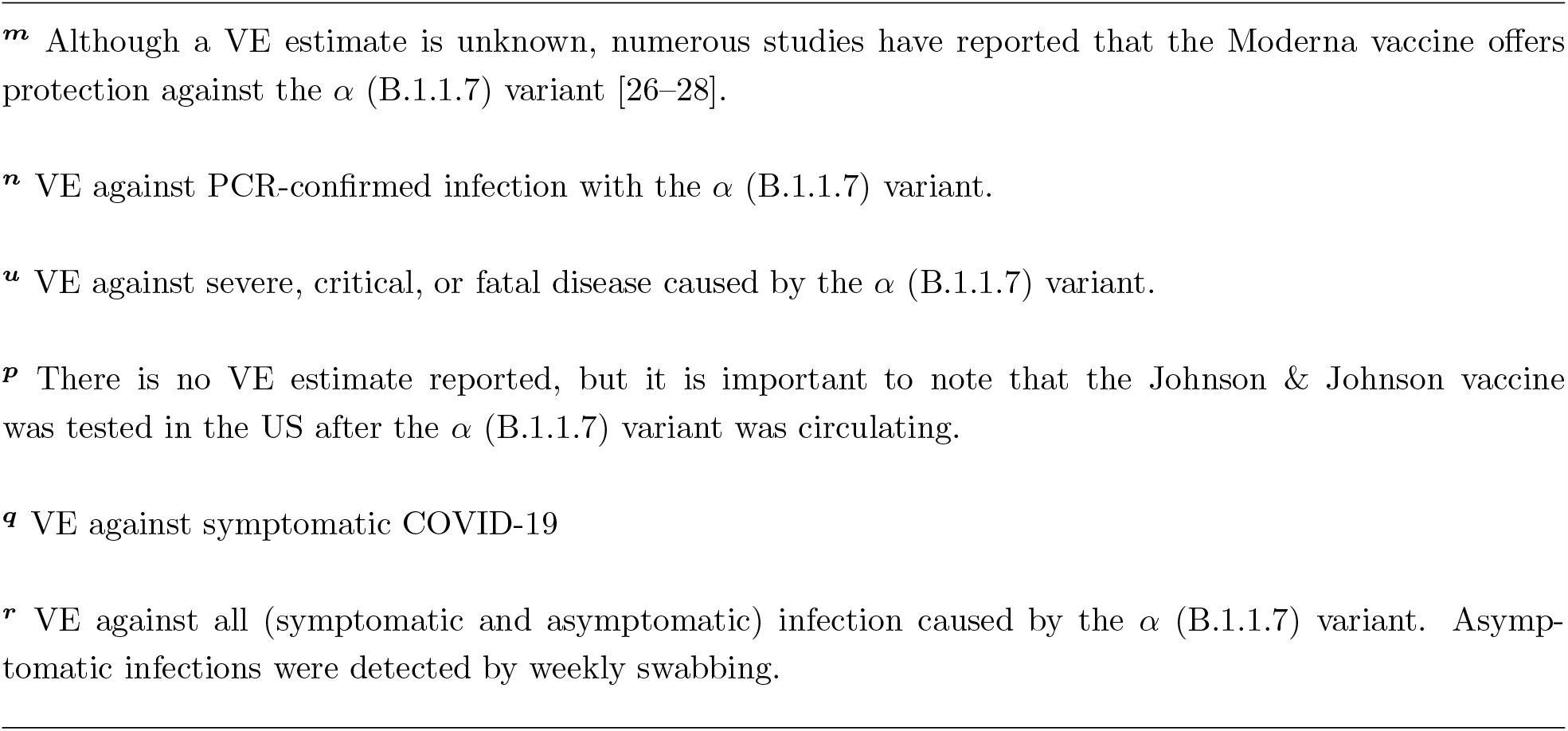
Vaccine efficacy against *α* (B.1.1.7) after dose 1.

| Company | Efficacy%, (95% CI),<br>time frame of estimate | References |
| --- | --- | --- |
| Moderna | Not reported<sup>m</sup> | [23] |
| Pfizer | 29.5, (22.9, 35.5)<sup>n</sup>, after<br>dose 1 | [23, 24] |
|  | 54.1, (26.1, 71.9)<sup>u</sup>, after<br>dose 1 | [24] |
|  | 49.2, (42.6, 55.0)<sup>q</sup>, after<br>dose 1 | [25] |
| Johnson & Johnson | Not reported<sup>p</sup> | [23] |
| AstraZeneca | 51.4, (47.3, 55.2)<sup>q</sup>, after<br>dose 1 | [25] |
| Novavax | 85.6<sup>*q</sup>, (CI not reported),<br>time frame not reported | [23, 26] |
| Sputnik V | Not reported | n/a |
| Sinovac | Not reported | n/a |
| Sinopharm | Not reported | n/a |

**Table 10:**
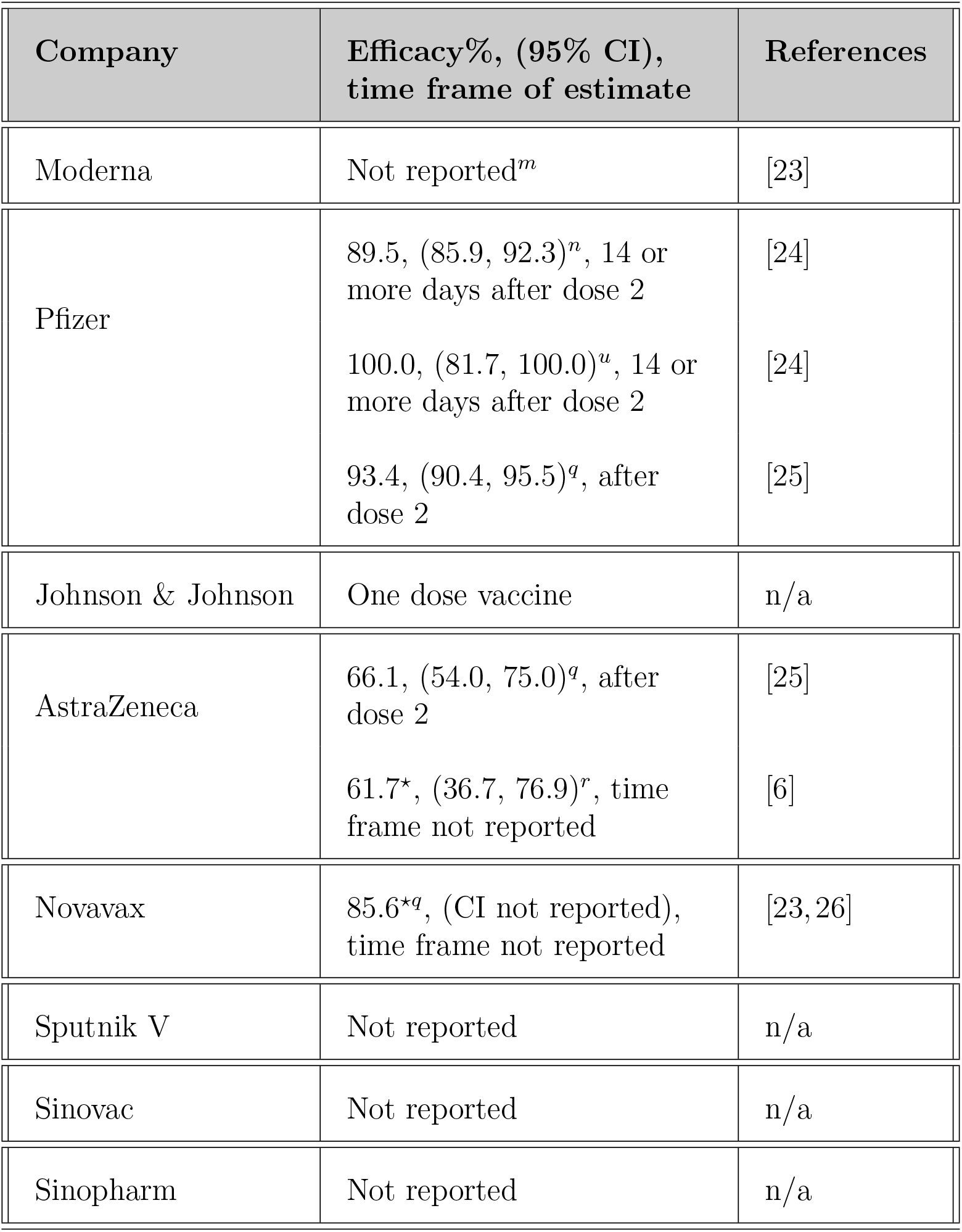
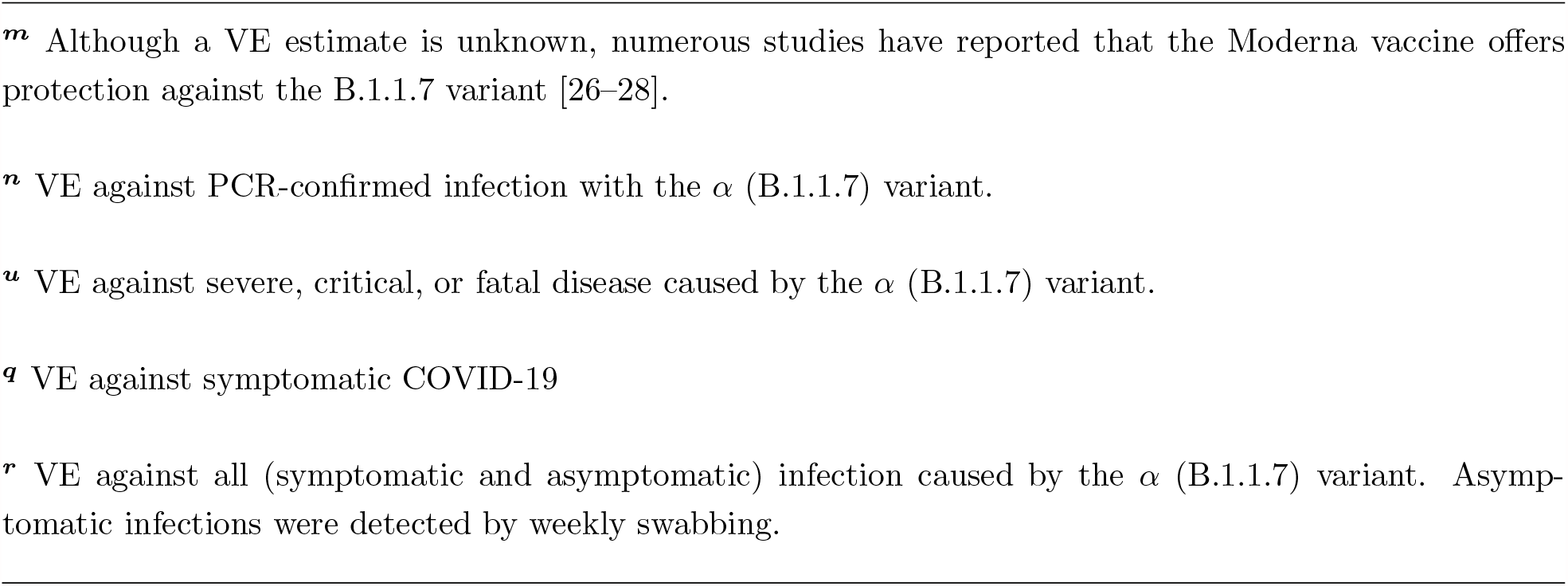
Vaccine efficacy against *α* (B.1.1.7) after dose 2.

| Company | Efficacy%, (95% CI),<br>time frame of estimate | References |
| --- | --- | --- |
| Moderna | Not reported<sup>m</sup> | [23] |
| Pfizer | 89.5, (85.9, 92.3)<sup>n</sup>, 14 or more days after dose 2 | [24] |
|  | 100.0, (81.7, 100.0)<sup>u</sup>, 14 or more days after dose 2 | [24] |
|  | 93.4, (90.4, 95.5)<sup>q</sup>, after dose 2 | [25] |
| Johnson & Johnson | One dose vaccine | n/a |
| AstraZeneca | 66.1, (54.0, 75.0)<sup>q</sup>, after dose 2 | [25] |
|  | 61.7<sup>*</sup>, (36.7, 76.9)<sup>r</sup>, time frame not reported | [6] |
| Novavax | 85.6<sup>*q</sup>, (CI not reported), time frame not reported | [23, 26] |
| Sputnik V | Not reported | n/a |
| Sinovac | Not reported | n/a |
| Sinopharm | Not reported | n/a |

**Table 11:** Vaccine efficacy against *γ* (P.1)

| Company | Efficacy%, (95% CI),<br>time frame of estimate | References |
| --- | --- | --- |
| Moderna | Not reported | n/a |
| Pfizer | Not reported | n/a |
| Johnson & Johnson | 66.2*, (51.0, 77.1)<sup>s</sup>, 14 or more days after vaccination | [12] |
|  | 68.1*, (48.8, 80.7)<sup>s</sup>, 28 or more days after vaccination | [12] |
| AstraZeneca | Not reported | n/a |
| Novavax | Not reported | n/a |
| Sputnik V | Not reported | n/a |
| Sinovac | 49.6, (11.3, 71.4)<sup>t</sup>, 14 or more days after dose 1 | [29] |
| Sinopharm | Not reported | n/a |

<sup>s</sup> VE against moderate to severe Covid-19 caused by the variant from the P.2 lineage carrying the E484K mutation.
<sup>t</sup> VE against symptomatic infection

**Table 12:**
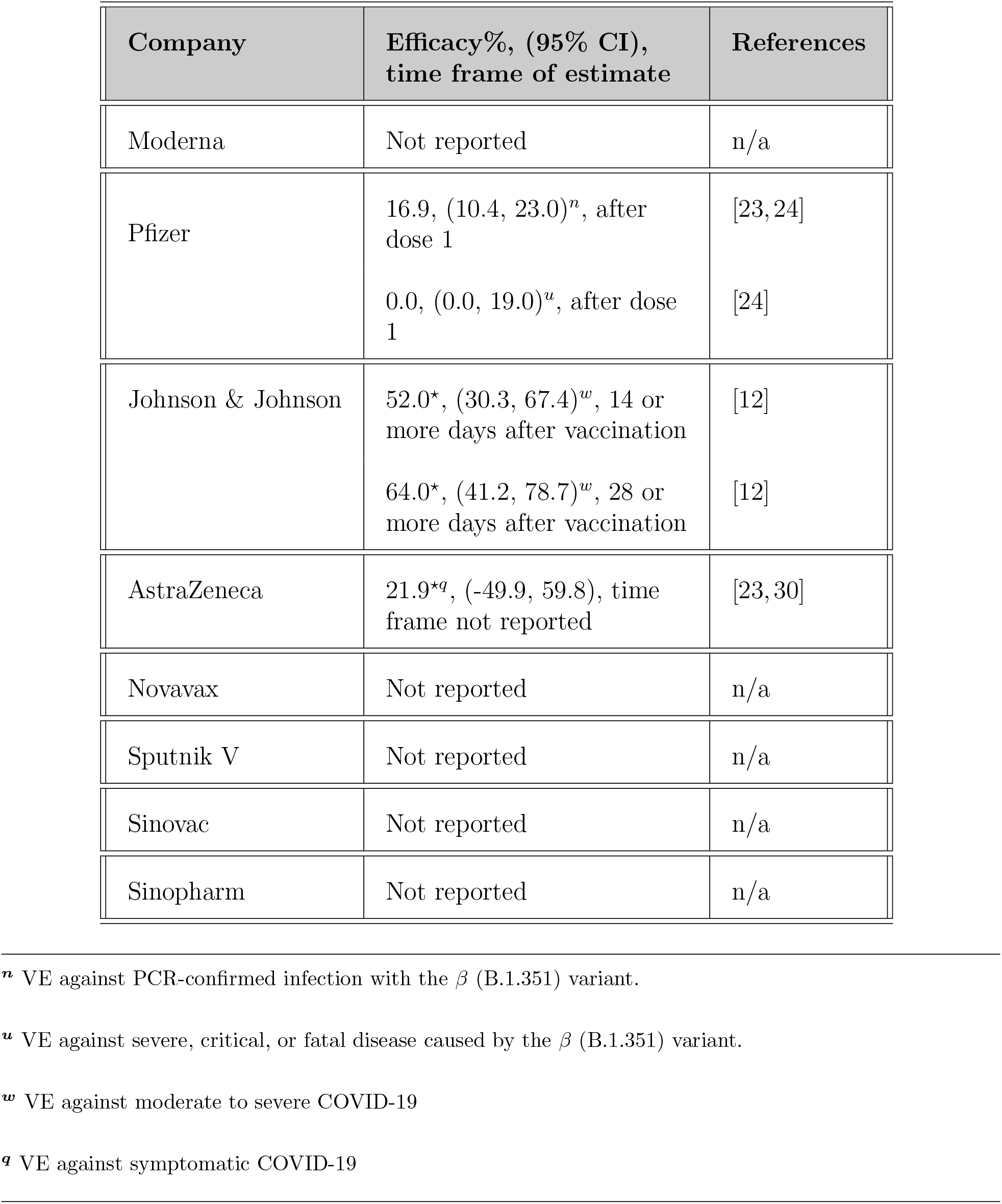
Vaccine efficacy against *β* (B.1.351) after dose 1.

| Company | Efficacy%, (95% CI),<br>time frame of estimate | References |
| --- | --- | --- |
| Moderna | Not reported | n/a |
| Pfizer | 16.9, (10.4, 23.0)<sup>n</sup>, after dose 1 | [23, 24] |
|  | 0.0, (0.0, 19.0)<sup>u</sup>, after dose 1 | [24] |
| Johnson & Johnson | 52.0*, (30.3, 67.4)<sup>w</sup>, 14 or more days after vaccination | [12] |
|  | 64.0*, (41.2, 78.7)<sup>w</sup>, 28 or more days after vaccination | [12] |
| AstraZeneca | 21.9<sup>*q</sup>, (-49.9, 59.8), time frame not reported | [23, 30] |
| Novavax | Not reported | n/a |
| Sputnik V | Not reported | n/a |
| Sinovac | Not reported | n/a |
| Sinopharm | Not reported | n/a |

**Table 13:** Vaccine efficacy against *β* (B.1.351) after dose 2.

| Company | Efficacy%, (95% CI),<br>time frame of estimate | References |
| --- | --- | --- |
| Moderna | Not reported | n/a |
| Pfizer | 75.0, (70.5, 78.9)<sup>n</sup>, 14 or<br>more days after dose 2 | [24] |
| Johnson & Johnson | One dose vaccine | n/a |
| AstraZeneca | 21.9*, (-49.9, 59.8)<sup>q</sup>, time<br>frame not reported | [23, 30] |
| Novavax | 49.4*, (6.1, 72.8)<sup>q</sup>, 7 days<br>after dose 2 | [23, 31] |
| Sputnik V | Not reported | n/a |
| Sinovac | Not reported | n/a |
| Sinopharm | Not reported | n/a |

**Table 14:**
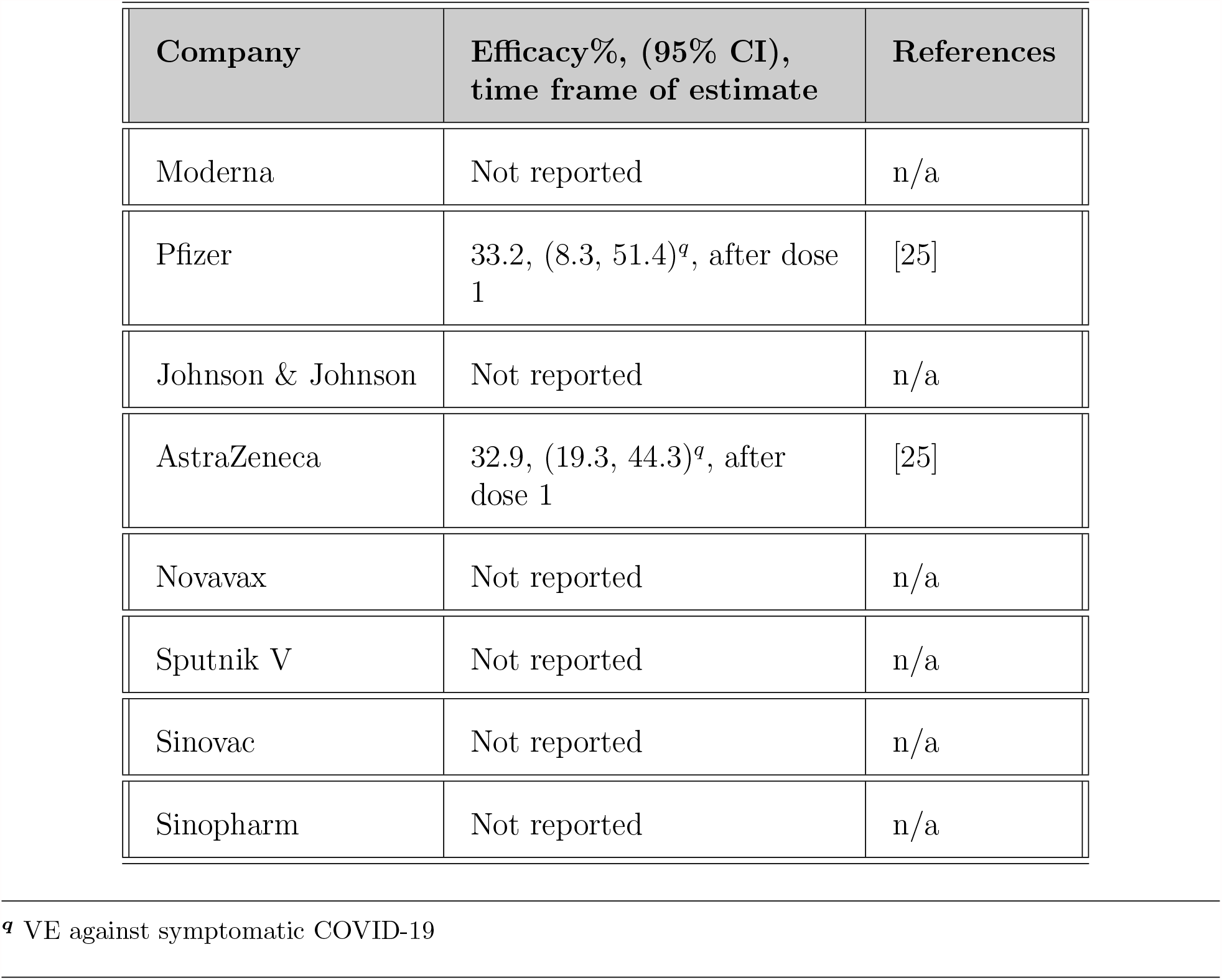
Vaccine efficacy against *δ* (B.1.617.2) after dose 1.

**Table 15:** Vaccine efficacy against *δ* (B.1.617.2) after dose 2.

| Company | Efficacy%, (95% CI),<br>time frame of estimate | References |
| --- | --- | --- |
| Moderna | Not reported | n/a |
| Pfizer | 87.9, (78.2, 93.2)<sup>q</sup>, after<br>dose 2 | [25] |
| Johnson & Johnson | Not reported | n/a |
| AstraZeneca | 59.8, (28.9, 77.3)<sup>q</sup>, after<br>dose 2 | [25] |
| Novavax | Not reported | n/a |
| Sputnik V | Not reported | n/a |
| Sinovac | Not reported | n/a |
| Sinopharm | Not reported | n/a |

**Table 16:** Viral neutralization of the variants of concern as compared with pre-existing variants.

| Company | Variant of concern | Neutralization by pseudovirion or live viral plaque assay | References |
| --- | --- | --- | --- |
| Moderna | B.1.1.7 ($\alpha$) | Decrease by 1.8x | [23] |
| | P.1 ($\gamma$) | Decrease by 4.5x | [23] |
| | B.1.351 ($\beta$) | Decrease by $\leq 8.6x$ | [23] |
| | B.1.617.2 ($\delta$) | Not reported | n/a |
| Pfizer | B.1.1.7 ($\alpha$) | Decrease by 2x | [23] |
| | P.1 ($\gamma$) | Decrease by 6.7x | [32] |
| | B.1.351 ($\beta$) | Decrease by $\leq 6.5x$ | [23] |
| | B.1.617.2 ($\delta$) | Not reported | n/a |
| Johnson & Johnson | B.1.1.7 ($\alpha$) | Not reported | n/a |
| | P.1 ($\gamma$) | Not reported | n/a |
| | B.1.351 ($\beta$) | Not reported | n/a |
| | B.1.617.2 ($\delta$) | Not reported | n/a |
| AstraZeneca | B.1.1.7 ($\alpha$) | Decrease by 9x | [6] |
| | P.1 ($\gamma$) | Not reported | n/a |
| | B.1.351 ($\beta$) | Decrease by $\leq 86\%$ | [23] |
| | B.1.617.2 ($\delta$) | Not reported | n/a |
| Novavax | B.1.1.7 ($\alpha$) | Decrease by 1.8x | [23] |
| | P.1 ($\gamma$) | Not reported | n/a |
| | B.1.351 ($\beta$) | Not reported | n/a |
| | B.1.617.2 ($\delta$) | Not reported | n/a |
| Sputnik V | B.1.1.7 ($\alpha$) | Not reported | n/a |
| | P.1 ($\gamma$) | Not reported | n/a |
| | B.1.351 ($\beta$) | Not reported | n/a |
| | B.1.617.2 ($\delta$) | Not reported | n/a |
| Sinovac | B.1.1.7 ($\alpha$) | Not reported | n/a |
| | P.1 ($\gamma$) | Not reported | n/a |
| | B.1.351 ($\beta$) | Not reported | n/a |
| | B.1.617.2 ($\delta$) | Not reported | n/a |
| Sinopharm | B.1.1.7 ($\alpha$) | Not reported | n/a |
| | P.1 ($\gamma$) | Not reported | n/a |
| | B.1.351 ($\beta$) | Not reported | n/a |
| | B.1.617.2 ($\delta$) | Not reported | n/a |

